# Tixagevimab/cilgavimab pre-exposure prophylaxis is associated with lower breakthrough infection risk in vaccinated solid organ transplant recipients during the Omicron wave

**DOI:** 10.1101/2022.05.17.22274980

**Authors:** Ayman Al Jurdi, Leela Morena, Mariesa Cote, Emily Bethea, Jamil Azzi, Leonardo V. Riella

## Abstract

The neutralizing monoclonal antibody combination of tixagevimab/cilgavimab has been shown to reduce the risk of SARS-CoV-2 infection in unvaccinated individuals during the Alpha (B.1.1.7) and Delta (B.1.617.2) waves. However, data on efficacy and safety of tixagevimab/cilgavimab in vaccinated solid organ transplant recipients during the Omicron wave is limited. To address this, we conducted a retrospective cohort study comparing 222 solid organ transplant recipients who received tixagevimab/cilgavimab for pre-exposure prophylaxis and 222 age-matched vaccinated solid organ transplant recipients who did not receive tixagevimab/cilgavimab. Subjects were followed for a mean of 67 ± 18 days. Kaplan-Meier estimates of the 60-day incidence of breakthrough infection were 1.8% in the tixagevimab/cilgavimab group and 4.7% in the control group (*P* = 0.045). Adverse events were uncommon, occurring in 4% of our cohort and most were mild. There was no significant change in serum creatinine or liver chemistries in kidney and liver transplant recipients respectively. In conclusion, we found that tixagevimab/cilgavimab use is safe and associated with a lower risk of breakthrough SARS-CoV-2 infection in vaccinated solid organ transplant recipients during the Omicron wave.

## 1. Introduction

Severe acute respiratory syndrome coronavirus 2 (SARS-CoV-2) infection in solid organ transplant recipients (SOTRs) is associated with higher mortality compared to immunocompetent individuals.^1^ Since antiviral responses to SARS-CoV-2 vaccines in SOTRs are attenuated,^2^ additional strategies such as monoclonal antibody pre-exposure prophylaxis has been developed.^3^ Tixagevimab and cilgavimab are neutralizing monoclonal antibodies directed against different epitopes of the receptor-binding domain of SARS-CoV-2 spike protein that have been associated with a lower risk of SARS-CoV-2 infection when used for pre-exposure prophylaxis in unvaccinated individuals.^4^ Based on that, tixagevimab/cilgavimab received emergency use authorization from the US Food and Drug Administration for pre-exposure prophylaxis against SARS-CoV-2 infection in high-risk populations.^5^ However, post-hoc analysis of the PROVENT trial showed a slightly higher incidence of serious cardiovascular events in the tixagevimab/cilgavimab.^5^ Furthermore, data on the safety and efficacy of tixagevimab/cilgavimab in SOTRs during the Omicron wave is limited as only a small number of SOTRs were included in the trial, all were unvaccinated, and the trial was performed during the period when the Alpha (B.1.1.7) and Delta (B.1.617.2) variants were prevalent.^4^

## 2. Materials and methods

### 2.1 Study design

To evaluate the safety and efficacy of tixagevimab/cilgavimab in SOTRs in a real-world setting during the Omicron period, we conducted a retrospective multicenter cohort study of kidney, liver and lung transplant recipients at our institutions who received tixagevimab/cilgavimab for SARS-CoV-2 pre-exposure prophylaxis. The primary outcome was the development of breakthrough SARS-CoV-2 infection, defined as a newly positive polymerase chain reaction or antigen test. Secondary outcomes included changes in allograft function and occurrence of adverse events after receiving tixagevimab/cilgavimab. The Mass General Brigham Research Patient Data Registry was used to identify a control group of age-matched vaccinated SOTRs who did not receive tixagevimab/cilgavimab for comparison. Follow-up for each patient in the control group was started on the same day that their age-matched counterpart in the tixagevimab/cilgavimab group received tixagevimab/cilgavimab to control for the variation in disease incidence. The study was approved by the Mass General Brigham institutional review board (protocol numbers: 2021P001235, 2019P002526 and 2017P000336).

## 2.2 Statistical analysis

Continuous variables are listed as mean ± standard deviation or median (interquartile range, IQR) depending on distribution. Categorical variables are listed as counts or percentages. Differences between paired sample continuous variables were assessed using a paired t test or Wilcoxon matched-pairs signed rank test depending on distribution. Kaplan-Meier curves were used to estimate the incidence of SARS-CoV-2 infection in the tixagevimab/cilgavimab and control groups with differences assessed using the log-rank test.

## 3. Results

### 3.1 Patient characteristics

222 solid organ transplant recipients were included in the study (Figure 1A). The median age was 65 years (IQR 55-72) and 39% were female. 25% had a history of coronary artery disease and 23% had a history of heart failure. 7% had a history of prior SARS-CoV-2 infection and 99% had received at least one dose of SARS-CoV-2 vaccine (Figure 1B). The median time from transplantation to tixagevimab/cilgavimab administration was 3.8 years (IQR 1.9-8.2). All doses were given between December 28^th^, 2021 and April 13^th^, 2022 (Figure 1C). 40.5% received the 150-150 mg dose, 59.0% received the 300-300 mg dose and 0.5% received the 450-450 mg dose.

**Figure 1.**
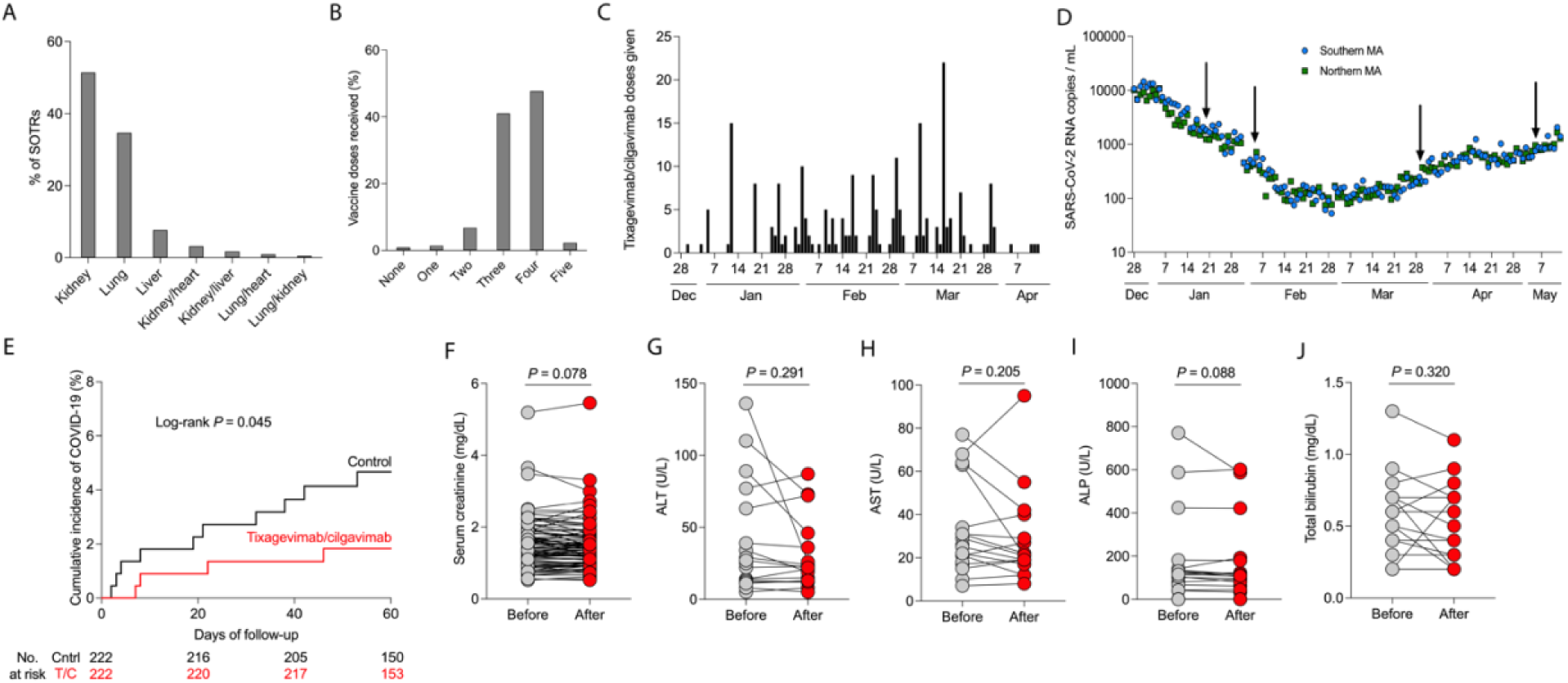
Tixagevimab/cilgavimab for pre-exposure prophylaxis in solid organ transplant recipients (SOTRs). (A) Types of SOTR subjects included in the study and (B) and the number of vaccines that subjects received. (C) Number of tixagevimab/cilgavimab doses given stratified by date of administration. (D) Wastewater SARS-CoV-RNA levels in Massachusetts (MA) by date. Data obtained from Massachusetts water resources authority (https://www.mwra.com/biobot/biobotdata.htm). Arrows indicate the dates of the four breakthrough infection events. (E) Survival curve of cumulative incidence of breakthrough coronarivus disease 19 (COVID-19) in SOTRs who received tixagevimab/cilgavimab (n=222) and age-matched vaccinated SOTRs (n=222). (F) Serum creatinine levels in kidney transplant recipients before and after tixagevimab/cilgavimab administration (n=92). (G) Alanine aminotransferase (ALT), (H) aspartate aminotransferase (AST), (I) alkaline phosphatase (ALP), and (J) total bilirubin levels in liver transplant recipients after tixagevimab/cilgavimab administration (n=16). (E) Statistic by log-rank test. (F) Statistic by paired t test. (G-J) Statistic by Wilcoxon matched-pairs signed rank test.

### 3.2 Breakthrough infection

At a mean follow-up of 67 ± 18 days after tixagevimab/cilgavimab, four SOTRs (1.8%) developed breakthrough infections (Figure 1D). Kaplan-Meier estimates of 60-day breakthrough infection rates were 1.8% in the tixagevimab/cilgavimab group and 4.7% in the age-matched vaccinated SOTR control group (*P* = 0.045, Figure 1E). In the tixagevimab/cilgavimab group, one subject was hospitalized and there were no deaths. In the control group, five were hospitalized and four died.

### 3.3 Safety and adverse events

Adverse events were uncommon, occurring only in 9 SOTRs (4%) at a median of 15 days (IQR 5-22) after tixagevimab/cilgavimab administration. The most common adverse events were nausea, vomiting or diarrhea (n=4, 1.8%), headache (n=3, 1.4%) and abdominal pain (n=2, 0.9%). Two patients (0.9%) developed new lung infiltrates with negative infectious evaluation, thought to be pneumonitis. In terms of cardiovascular adverse events, one patient (0.5%) developed a mild heart failure exacerbation and one (0.5%) developed new atrial fibrillation requiring cardioversion.

In kidney transplant recipients, there was no significant change in serum creatinine levels measured at a median of 30 days (IQR 20-46) after tixagevimab/cilgavimab compared to baseline (Figure 1F, *P* = 0.078). In liver transplant recipients, there was no significant change in alanine aminotransferase (Figure 1G, *P* = 0.291), aspartate aminotransferase (Figure 1H, *P* = 0.205), alkaline phosphatase (Figure 1I, *P* = 0.088) or total bilirubin levels (Figure 1J, *P* = 0.320) measured at a median of 22 days (IQR 12-43) after tixagevimab/cilgavimab.

## 4 Discussion

In this study, we report our early experience with using tixagevimab/cilgavimab for pre-exposure prophylaxis in SOTRs during the Omicron wave. We found that tixagevimab/cilgavimab use was associated with a significantly lower risk of SARS-CoV-2 breakthrough infection, compared to an age-matched vaccinated control group of SOTRs at our centers. Hospitalizations and deaths from SARS-CoV-2 infection were also numerically lower in the tixagevimab/cilgavimab group compared to control, but the number of events was too small to evaluate differences between the two groups. We also found that tixagevimab/cilgavimab was safe with a low rate of adverse events and no change in allograft function in kidney or liver transplant recipients. Our study adds to the literature by including vaccinated SOTRs, which were not included in the original trial,^4^ and evaluating breakthrough infections during the Omicron period, as opposed to the Alpha and Delta period during which the original trial was conducted.^4^ The limitations of our study include its observational nature, retrospective design, lack of ability to detect asymptomatic breakthrough infections and possible differences between the tixagevimab/cilgavimab and control groups that could have influenced the rate of breakthrough infection. Despite these limitations, our study adds evidence of the efficacy and safety of tixagevimab/cilgavimab for pre-exposure prophylaxis in vaccinated SOTRs during the Omicron wave.

## Supporting information

Disclosures

## Data Availability

Data to support the findings in the study are available from the corresponding author upon request.

## Abbreviations

IQR: interquartile range
SARS-CoV-2: Severe acute respiratory syndrome coronavirus 2
SOTR: Solid organ transplant recipient

## Acknowledgments

None.

## Data availability statement

Data to support the findings in the study are available from the corresponding author upon request.

## References

1. Caillard S, Chavarot N, Francois H, et al. Is COVID-19 infection more severe in kidney transplant recipients? Am J Transplant. 2021;21(3):1295–1303. doi:10.1111/ajt.16424

2. Al Jurdi A, Gassen RB, Borges TJ, et al. Non-Invasive Monitoring for Rejection in Kidney Transplant Recipients After SARS-CoV-2 mRNA Vaccination. Front Immunol. 2022;13:838985. doi:10.3389/fimmu.2022.838985

3. Loo Y-M, McTamney PM, Arends RH, et al. The SARS-CoV-2 monoclonal antibody combination, AZD7442, is protective in nonhuman primates and has an extended half-life in humans. Sci Transl Med. 2022;14(635):eabl8124. doi:10.1126/scitranslmed.abl8124

4. Levin MJ, Ustianowski A, De Wit S, et al. Intramuscular AZD7442 (Tixagevimab-Cilgavimab) for Prevention of Covid-19. N Engl J Med. Published online April 20, 2022. doi:10.1056/NEJMoa2116620

5. Tixagevimab and Cilgavimab (Evusheld) for Pre-Exposure Prophylaxis of COVID-19. JAMA. 2022;327(4):384–385. doi:10.1001/jama.2021.24931

